# Stressful life events recorded in 39.7 million clinical notes: associations with mental disorders, comorbidity, and frailty

**DOI:** 10.64898/2026.11.06.26367427

**Authors:** Anna A. Peltola, Esha Khan, Juho Kaijansinkko, Nina Alexander, Anna Tirkkonen, Mikaela B. von Bonsdorff, Jake Lin, Juulia Jylhävä

## Abstract

**Background:** Despite being harmful to both mental and physical health, stressful life events are not recorded in structured electronic health records. They appear sporadically in narrative clinical notes, where they are difficult to analyze at scale. Natural language processing (NLP) has made clinical notes more available for research, but applications to stressful life events remain limited, leaving their documentation patterns and prognostic value understudied. By applying NLP, we identified stressful life events from clinical notes and examined their associations with mental disorders, comorbidity, and frailty.

**Methods:** We analyzed 39.7 million clinical notes, recorded between 2010 and 2023, from 202,988 patients in Central Finland Wellbeing Services County. NLP was applied to identify stressful life events in five categories: grief, divorce, financial distress, violence, and family problems. Associations with the risk of mental disorders (stress-related disorders, anxiety, and depression) were estimated using time-varying Cox proportional hazards models, and associations with physical health outcomes (comorbidity and frailty) using generalized linear mixed models.

**Results:** Stressful life events were identified from clinical notes with high accuracy (F1 scores 0.81–0.95). Event prevalences ranged from 0.5% for family problems to 22.1% for grief. Most individual events were associated with mental disorders, comorbidity, and frailty. The number of different life events was associated with a higher risk of stress-related (hazard ratio [HR], 1.62; 95% confidence interval [CI], 1.58–1.67), anxiety (HR, 2.08; 95% CI, 2.03–2.14), and depressive disorders (HR, 1.99; 95% CI, 1.94–2.03), and with higher comorbidity (rate ratio, 1.58; 95% CI,1.56–1.59) and frailty (mean ratio, 1.57; 95% CI, 1.56–1.58).

**Conclusions:** Stressful life events were accurately identified from clinical notes and were associated with a higher risk of mental disorders, comorbidity, and frailty. Even so, violence and financial distress, the two events with comparable national estimates, were documented substantially less often than those estimates, suggesting underrecording of stressful life events in clinical settings.

## 1. Introduction

Despite being harmful to both mental and physical health (1–6), stressful life events are not recorded routinely in structured electronic health records. Instead, they are documented sporadically within narrative clinical notes. Because these unstructured notes are difficult to analyze at scale (7,8), the extent to which stressful life events are reported in clinical settings and whether the reported events have prognostic relevance remains understudied.

Natural language processing (NLP) has made clinical notes more accessible for research purposes, but applications to stressful life events remain preliminary. A recent scoping review found only a few studies that identified life events from clinical notes (7). Some of these studies used the identified events to predict psychiatric outcomes, such as suicide risk (9), but none examined physical health outcomes.

For physical health, evidence from survey-based cohorts suggests stressful life events are associated with comorbidity and biological aging markers (1–6). Some of these outcomes can also be derived from diagnostic codes in electronic health records (10), allowing exposure and outcome to be observed within the same patient history. Electronic health records also capture outcomes that surveys reach unevenly, such as frailty – a measure of accumulated health deficits that requires detailed clinical information (11).

In this study, we identify stressful life events from 39.7 million Finnish-language clinical notes from the Central Finland Wellbeing Services County. We identify five categories of life events – grief, divorce, financial distress, violence, and family problems – and examine their associations with common mental disorders, comorbidity, and frailty. To our knowledge, this is the first life event extraction study in a non-English setting and the first to relate extracted life events to physical health outcomes.

## 2. Methods

### 2.1 Study population

The Central Finland Wellbeing Services County provides primary, secondary, and tertiary public healthcare for all residents in the region. Our study population comprises all individuals aged 34 and above with at least one clinical encounter documented in electronic health records between 2010 and 2023. These records include 39,739,126 clinical notes from 202,988 patients, comprising all free-text documentation by healthcare professionals – progress notes, admission and discharge summaries, nursing notes, psychiatric notes, assessments of social conditions, and patient and family communication.

### 2.2 Analytical sample

In the primary analytical sample, we included 177,028 individuals with at least one healthcare visit and no baseline psychiatric diagnoses. Individual follow-up was defined as the time between the first and last documented healthcare visit between 2010 and 2023.

### 2.3 Stressful life events

By stressful life events, we refer to events that are broadly perceived as disruptive and undesirable (7). This general definition is suited to clinical notes, which do not allow us to assess how a patient experienced an event.

#### 2.3.1 Category definition

Five event categories were identified: grief, divorce, violence, family problems, and financial distress (Table 1). Four of these overlap with domains represented in the *Stress and Adversity Inventory for adults* (12), a comprehensive measure of lifetime stress. Family problems, as defined here, have no direct equivalent in existing survey instruments and instead emerged from reviewing the clinical notes.

**Table 1.** Stressful life event categories and their definitions.

| **Category** | **Definition** |
|---|---|
| **Grief** | Death of a close person (partner, child, parent, sibling, or friend), bereavement, widowhood |
| **Divorce** | Divorce or separation from any romantic relationship, regardless of formal marital status |
| **Financial distress** | Unemployment, financial difficulties, debt and debt enforcement proceedings, and receipt of last-resort social assistance |
| **Violence** | Physical, sexual, or emotional violence and abuse, including both direct and vicarious exposure – for instance, a parent learning their child has been sexually abused |
| **Family problems** | Contact with child protective or welfare services, child placement, custody disputes, and family crisis – primarily from a parental perspective |

The selection of all five was further guided by discriminability in clinical text – that is, sufficient frequency in clinical documentation and lexical and semantic distinctiveness between categories. Categories were kept broad enough to capture the variety of expressions encountered in clinical notes while remaining conceptually coherent.

The integration of Finnish healthcare and social services shaped the family problems category specifically. Family workers, counsellors, and child welfare services work closely alongside healthcare, and this inter-agency collaboration is routinely documented in clinical notes. Clinicians are also obligated to notify child protective services when they have concerns about a child’s welfare. This anchored the category to institutional language rather than more diffuse descriptions of family situations, which would have been difficult to extract reliably.

#### 2.3.2 Named entity recognition

To identify the five stressful life event categories in clinical notes, we used named entity recognition (NER) – a natural language processing technique for identifying predefined concepts in free text.

We adapted the pipeline developed by Lin et al. (13), which proceeded in three phases: (1) pre-filtering, (2) annotation, and (3) model training and inference.

##### Pre-filtering

For each category, we compiled a list of Finnish-language keywords and used these to search all clinical notes, retaining only those with at least one keyword match.

##### Annotation

We drew a random sample of 5,000 keyword-filtered notes per category. Reading through these notes allowed us to expand and refine the keyword lists by identifying unanticipated expressions and addressing recurring linguistic problems: terms with overlapping surface forms were combined into a single category (e.g., physical and emotional violence were captured as violence), and polysemous terms were disambiguated using contextual phrases. All identified expressions – both those matching keywords and unanticipated ones – were annotated, and notes without annotations were treated as absence of the event. Annotations were made and approved by separate authors.

##### Model training and inference

The annotated notes were used to fine-tune FinBERT (14), a Finnish-language transformer model, on GPU-enabled cloud infrastructure of the Helsinki and Uusimaa Hospital District. The dataset was split into training, validation, and test sets in 80/10/10 proportions, and hyperparameters were tuned iteratively with model selection based on F1 score.

The trained model was then applied to all keyword-filtered notes, and an inference token probability score of at least 0.95 was applied to reduce false positives. Further technical details follow Lin et al. (13).

#### 2.3.3 Model performance evaluation

Model performance was evaluated on the held-out test set – the 10% of annotated notes set aside for evaluation. Precision indicates the proportion of identified mentions that are true positives, recall indicates the proportion of true positives that were identified, and F1 score is their harmonic mean. Performance metrics were based on entity type matching, where an event mention identified by the model was considered correct if it overlapped with a manually annotated mention of the same category.

#### 2.3.4 Stressful life events as exposure variables

For the statistical analyses, each life event category was coded as a binary time-varying variable: 0 until the calendar year of the first documented mention and 1 from that year onward, so that a documented event remained an exposure for the rest of follow-up. As an additional exposure, we used the sum of life events, defined as the number of different categories documented up to a given year (0–5). The sum increased by one each year a new category was documented.

#### 2.3.5 Documentation of life events

We described the documentation of each life event category in two ways. First, we calculated the number and proportion of patients with the event documented at least once during follow-up. Second, we calculated the proportion of patients with a healthcare visit in a calendar year who had the event documented in that year, and averaged these proportions over 2010–2023.

### 2.4 Mental disorders as outcomes

#### 2.4.1 Stress-related disorders

Stress-related disorders were defined as the F43 category of ICD-10, which includes the diagnoses of acute stress reaction (F43.0), posttraumatic stress disorder (F43.1), adjustment disorders (F43.2), and other reactions to severe stress (F43.8).

#### 2.4.2 Anxiety disorders

Anxiety disorders were defined as ICD-10 categories F40–F41, which include phobic anxiety disorders (F40) and other anxiety disorders (F41), such as panic disorder (F41.0), generalized anxiety disorder (F41.1), and mixed anxiety and depressive disorder (F41.2).

#### 2.4.3 Depressive disorders

Depressive disorders were defined as ICD-10 categories F32–F33, which include depressive episodes (F32) and recurrent depressive disorder (F33).

### 2.5 Physical health outcomes

#### 2.5.1 Comorbidity

Comorbidity was defined by the Charlson comorbidity index (CCI) (15), applying the Swedish register-based adaptation described by Ludvigsson et al. (10). 18 ICD-10 diagnostic codes were assigned weights and summed into a continuous score. For each calendar year, the score included all conditions recorded from the individual’s baseline up to and including that year.

#### 2.5.2 Frailty

Frailty was defined using the Hospital frailty risk score (HFRS) (11), which assigns weights to 109 ICD-10 diagnoses and sums them into a continuous score. The score was updated at the end of each calendar year to include all codes recorded since the individual baseline. To restrict the score to physical health only, psychiatric and psychosocial codes (F10, F32, R45, Z60, Z73) were excluded. Weights for the remaining codes were unchanged.

### 2.6 Covariates

All models were adjusted for sex and age. Models for comorbidity and frailty were additionally adjusted for the corresponding baseline measure, and models for psychiatric outcomes for baseline comorbidity.

### 2.7 Statistical analyses

#### 2.7.1 Analyses for mental disorders

Associations with the risk of stress-related, anxiety and depressive disorders were estimated using Cox proportional hazards models with age as the time scale and standard errors clustered by individual. Patients were followed until the first diagnosis of the outcome or the end of their follow-up. Patients with a baseline diagnosis of any of the three disorders were excluded from all models.

For each outcome, univariable models included one life event at a time, or the sum of life events. Multivariable models included either the five life events simultaneously or the sum of life events. Univariable models were adjusted for sex, and multivariable models were additionally adjusted for baseline comorbidity.

The data were organized by calendar year, so the order of an event and a diagnosis in the same year was determined from the exact dates. Individuals whose event was first documented after the diagnosis date during the same year were excluded from the analysis of that outcome, because the risk of exposure misclassification was high: the event may have preceded the diagnosis and been recorded only afterwards. Events documented on the day of diagnosis were kept as exposures.

Predictive performance of the models was assessed with the Harrell’s C-index, both in the full sample and in a random 70/30 split of participants into training and test sets. The models were fitted in the training set and evaluated in the test set.

#### 2.7.2 Analyses for comorbidity and frailty

Associations with comorbidity and frailty were estimated using generalized linear mixed models. The models included random intercepts and random slopes for age, so that individuals differed in both their level and their rate of change with age. Comorbidity, a right-skewed integer score, was modelled using a Poisson distribution, and frailty, a right-skewed continuous score with many zeros, using a Tweedie distribution. All models used a log link, so the exponentiated coefficients are rate ratios for comorbidity and mean ratios for frailty. For each outcome, univariable models included one life event at a time, or the sum of life events. Multivariable models included either the five life events simultaneously or the sum of life events. Univariable models were adjusted for centered age and sex, and multivariable models were additionally adjusted for baseline value of the outcome.

#### 2.7.3 Moderation by mental disorders

To examine whether the associations for physical health outcomes differed by psychiatric diagnosis, we added an interaction between each life event and any psychiatric diagnosis (stress-related, anxiety or depressive disorder) to the mixed models. Psychiatric diagnosis was a time-varying variable, coded 0 before and 1 from the date of the first diagnosis of a stress-related, anxiety or depressive disorder. As in the models for psychiatric outcomes, individuals whose event was first documented after the diagnosis date in the same year were excluded to avoid misclassification.

### 2.8 Infrastructure

All data were stored and analyzed within Helsinki and Uusimaa Hospital District’s secure server. Analyses were performed using R version 4.6.2, with packages including *survival, glmmTMB, tidyverse*, and *emmeans*. Statistical significance was set at a two-sided α of 0.05.

## 3. Results

### 3.1 Model performance metrics

Stressful life events were identified from clinical notes with high accuracy. F1 scores ranged from 0.81 for grief and family problems to 0.95 for violence (Table 2). Precision ranged from 0.70 for grief to 0.92 for violence, and recall from 0.79 for family problems to 0.98 for violence. Most frequent expressions identified for each category are listed in eTable 1.

**Table 2.** Performance metrics for each category in the held-out test set.

| **Category** | **F1** | **Precision** | **Recall** |
|---|---|---|---|
| **Grief** | 0.81 | 0.70 | 0.95 |
| **Divorce** | 0.91 | 0.87 | 0.95 |
| **Financial distress** | 0.88 | 0.88 | 0.88 |
| **Violence** | 0.95 | 0.92 | 0.98 |
| **Family problems** | 0.81 | 0.83 | 0.79 |

### 3.2 Descriptive results

The analytical sample included 177,028 patients (51.2% female) with at least one healthcare visit and no baseline psychiatric diagnosis (Table 3). At baseline, patients were aged 59.4±13.5 years and were followed for 9.4±4.9 years. The median comorbidity and frailty scores were 0 (IQR 0–0).

**Table 3.** Characteristics of the cohort and documented life events. Any life event documented means at least one of the five categories was documented during follow-up. SD = standard deviation; IQR = interquartile range.

|  | All | Any life event documented | None documented |
|---|---|---|---|
| Patients, n | 177,028 | 52,209 | 124,819 |
| Follow-up in years, mean (SD) | 9.4 (4.9) | 11.6 (3.5) | 8.4 (5.2) |
| **Sex, n (%)** |  |  |  |
| Female | 90,705 (51.2) | 30,260 (58.0) | 60,445 (48.4) |
| Male | 86,323 (48.8) | 21,949 (42.0) | 64,374 (51.6) |
| Age at baseline in years, mean (SD) | 59.4 (13.5) | 59.8 (13.8) | 59.2 (13.3) |
| Comorbidity at baseline, median (IQR) | 0 (0–0) | 0 (0–0) | 0 (0–0) |
| Frailty at baseline, median (IQR) | 0 (0–0) | 0 (0–0) | 0 (0–0) |
| **Life events documented, n (%)** |  |  |  |
| Grief | 39,164 (22.1) | 39,164 (75.0) | . |
| Divorce | 8,757 (4.9) | 8,757 (16.8) | . |
| Financial distress | 18,969 (10.7) | 18,969 (36.3) | . |
| Violence | 3,420 (1.9) | 3,420 (6.6) | . |
| Family problems | 956 (0.5) | 956 (1.8) | . |
| **Number of life event categories, n (%)** |  |  |  |
| 0 | 124,819 (70.5) | . | . |
| 1 | 38,166 (21.6) | 38,166 (73.1) | . |
| 2 | 10,093 (5.7) | 10,093 (19.3) | . |
| 3 or more | 3,950 (2.2) | 3,950 (7.6) | . |

At least one life event was documented in 52,209 patients (29.5%). One life event category was documented in 38,166 patients (21.6%), two in 10,093 (5.7%) and three or more in 3,950 (2.2%). Grief was the most frequently documented life event (22.1%), followed by financial distress (10.7%), divorce (4.9%), violence (1.9%) and family problems (0.5%). In a given year, the proportion of patients with a visit who had an event documented that year was 4.7% for grief, 2.7% for financial distress, 1.0% for divorce, 0.4% for violence and 0.1% for family problems.

Compared with patients without a documented life event, those with any documented life event had a longer follow-up (11.6±3.5 versus 8.4±5.2 years) and were more likely to be female (58.0% versus 48.4%). Baseline age (59.8±13.8 versus 59.2±13.3 years), comorbidity (0 [0-0] versus 0 [0-0]), and frailty (0 [0-0] versus 0 [0-0]) were similar in both groups.

### 3.3 Results for mental disorders

In univariable models (eTable 2), each life event and the sum of life events were associated with a higher risk of stress-related, anxiety and depressive disorders (all *p* < 0.001). When all life events were entered in a mutually adjusted model (Table 4), the associations remained significant except those of family problems with anxiety and depressive disorders. The strongest association was observed between divorce and depressive disorders (hazard ratio [HR] 2.66, 95% confidence interval [CI] 2.43–2.91).

**Table 4.**
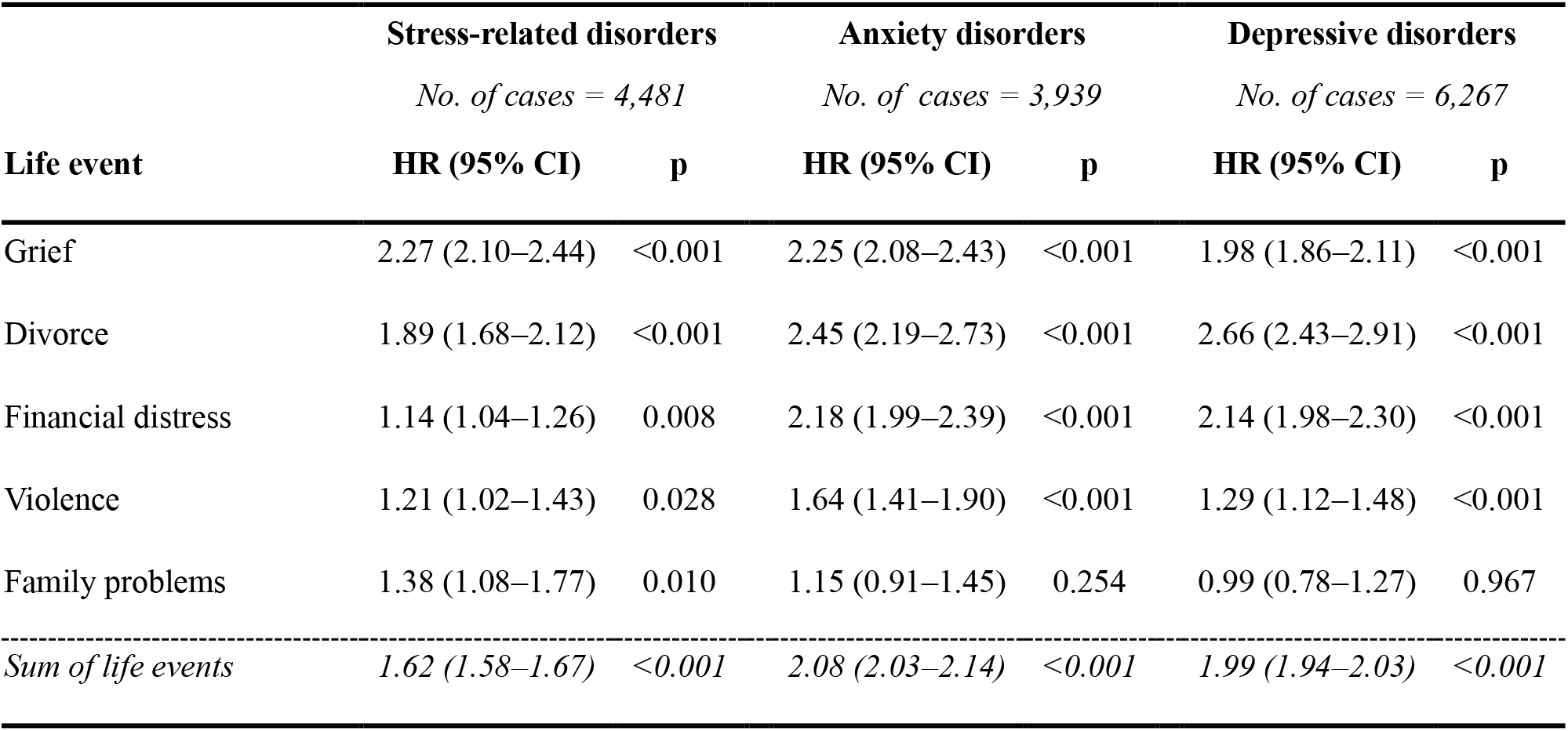
Life events and risk of stress-related, anxiety and depressive disorders. Results from time-varying Cox proportional hazards models with age as the time scale, adjusted for sex and baseline CCI. The five life event categories are mutually adjusted; the sum of life events was estimated in a separate model. Abbreviations: HR = hazard ratio; CI = 95% confidence interval.

|  | **Stress-related disorders** |  | **Anxiety disorders** |  | **Depressive disorders** |  |
|---|---|---|---|---|---|---|
|  | *No. of cases = 4,481* |  | *No. of cases = 3,939* |  | *No. of cases = 6,267* |  |
| **Life event** | **HR (95% CI)** | **p** | **HR (95% CI)** | **p** | **HR (95% CI)** | **p** |
| Grief | 2.27 (2.10–2.44) | <0.001 | 2.25 (2.08–2.43) | <0.001 | 1.98 (1.86–2.11) | <0.001 |
| Divorce | 1.89 (1.68–2.12) | <0.001 | 2.45 (2.19–2.73) | <0.001 | 2.66 (2.43–2.91) | <0.001 |
| Financial distress | 1.14 (1.04–1.26) | 0.008 | 2.18 (1.99–2.39) | <0.001 | 2.14 (1.98–2.30) | <0.001 |
| Violence | 1.21 (1.02–1.43) | 0.028 | 1.64 (1.41–1.90) | <0.001 | 1.29 (1.12–1.48) | <0.001 |
| Family problems | 1.38 (1.08–1.77) | 0.010 | 1.15 (0.91–1.45) | 0.254 | 0.99 (0.78–1.27) | 0.967 |
| *Sum of life events* | *1.62 (1.58–1.67)* | *<0.001* | *2.08 (2.03–2.14)* | *<0.001* | *1.99 (1.94–2.03)* | *<0.001* |

Each additional life event was associated with a higher risk of stress-related disorders (HR 1.62, 95% CI 1.58–1.67), anxiety disorders (HR 2.08, 95% CI 2.03–2.14) and depressive disorders (HR 1.99, 95% CI 1.94–2.03). Associations were stronger for anxiety and depressive disorders than for stress-related disorders. Concordance of the multivariable models was 0.68, 0.70 and 0.68, respectively (eTable 3), and was similar in the training and test sets (eTable 4).

Results were largely similar in a sensitivity analysis, where individuals with life events documented on the date of diagnosis were additionally excluded (eTable 5). The exceptions were grief, for which the hazard ratios were lower for all three outcomes (HR 1.53–2.05), and violence, which was no longer associated with stress-related disorders (HR 1.13, 95% CI 0.93–1.37).

### 3.4 Results for comorbidity and frailty

In univariable models (eTable 6), all life events and the sum of life events were associated with higher comorbidity and frailty (all *p* < 0.001). The associations remained significant in the mutually adjusted models (Table 5). The strongest associations were observed for financial distress, with a rate ratio (RR) of 1.76 (95% confidence interval [CI] 1.73–1.79) for comorbidity and a mean ratio (MR) of 1.70 (95% CI 1.68–1.73) for frailty.

**Table 5.** Life events and trajectories of comorbidity and frailty. Results from generalised linear mixed models with a random intercept and a random slope for age, fitted as Poisson for comorbidity and Tweedie for frailty. Comorbidity was measured with the Charlson comorbidity index (CCI) and frailty with the Hospital frailty risk score (HFRS). Each model is adjusted for sex and the baseline value of the respective outcome. The five life event categories are mutually adjusted; the sum of life events was estimated in a separate model. CI = 95% confidence interval.

|  | **Comorbidity** |  | **Frailty** |  |
|---|---|---|---|---|
| **Life event** | **Rate ratio (95% CI)** | **p** | **Mean ratio (95% CI)** | **p** |
| Grief | 1.54 (1.52–1.55) | <0.001 | 1.54 (1.53–1.55) | <0.001 |
| Divorce | 1.54 (1.49–1.58) | <0.001 | 1.58 (1.55–1.61) | <0.001 |
| Financial distress | 1.76 (1.73–1.79) | <0.001 | 1.70 (1.68–1.73) | <0.001 |
| Violence | 1.34 (1.28–1.40) | <0.001 | 1.38 (1.34–1.41) | <0.001 |
| Family problems | 1.31 (1.20–1.43) | <0.001 | 1.40 (1.33–1.48) | <0.001 |
| *Sum of life events* | *1.58 (1.56–1.59)* | *<0.001* | *1.57 (1.56–1.58)* | *<0.001* |

Each additional life event was associated with an RR of 1.58 (95% CI 1.56–1.59) for comorbidity and an MR of 1.57 (95% CI 1.56–1.58) for frailty. Marginal R^2^ was 0.38 for comorbidity and 0.39 for frailty, in both the models with individual life events and those with the sum of life events (eTable 7).

### 3.5 Moderation by mental disorders

The associations of life events with comorbidity and frailty differed by psychiatric diagnosis (Table 6). Compared with individuals without a psychiatric diagnosis, those with a diagnosis had weaker associations of grief, divorce and financial distress with both outcomes, and of violence with frailty (*p* for interaction < 0.001 for all). There was no evidence of interaction for violence and family problems with comorbidity or for family problems with frailty. For any documented life event versus none, the RR for comorbidity was 1.76 (95% CI 1.74–1.78) without and 1.33 (95% CI 1.29– 1.38) with a psychiatric diagnosis, and the MR for frailty was 1.80 (95% CI 1.79–1.82) and 1.35 (95% CI 1.32–1.38), respectively (*p* for interaction < 0.001 for both).

**Table 6.** Associations between life events and comorbidity and frailty by psychiatric diagnosis. Marginal mean ratios (95% CI) for each life event among people without and with a psychiatric diagnosis, averaged over sex and evaluated at the mean age. p = p-value for the difference in marginal mean ratios by psychiatric diagnosis. Comorbidity was measured with the Charlson comorbidity index and frailty with the Hospital frailty risk score.

|  | **Comorbidity** |  |  | **Frailty** |  |  |
|---|---|---|---|---|---|---|
| **Life event** | **No psychiatric diagnosis** | **Psychiatric diagnosis** | **p** | **No psychiatric diagnosis** | **Psychiatric diagnosis** | **p** |
| Grief | 1.61 (1.59–1.63) | 1.21 (1.17–1.25) | <0.001 | 1.60 (1.59–1.62) | 1.19 (1.17–1.22) | <0.001 |
| Divorce | 1.77 (1.72–1.83) | 1.50 (1.43–1.58) | <0.001 | 1.81 (1.77–1.84) | 1.42 (1.37–1.46) | <0.001 |
| Financial distress | 1.92 (1.88–1.96) | 1.56 (1.50–1.62) | <0.001 | 1.85 (1.83–1.88) | 1.44 (1.41–1.48) | <0.001 |
| Violence | 1.56 (1.48–1.65) | 1.46 (1.34–1.58) | 0.104 | 1.59 (1.55–1.64) | 1.31 (1.25–1.36) | <0.001 |
| Family problems | 1.79 (1.58–2.03) | 1.57 (1.37–1.80) | 0.120 | 1.68 (1.56–1.80) | 1.66 (1.53–1.81) | 0.881 |
| *Sum of life events\** | *1.76 (1.74–1.78)* | *1.33 (1.29–1.38)* | *<0.001* | *1.80 (1.79–1.82)* | *1.35 (1.32–1.38)* | *<0.001* |

\* 1 or more versus 0 life events (ref.).

## 4. Discussion

Even when sparsely recorded, stressful life events were associated with a higher risk of mental disorders and with higher comorbidity and frailty. The prevalence of events ranged from 0.5% (family problems) to 22.1% (grief), and each additional event category was associated with a higher risk of stress-related disorders (hazard ratio [HR] 1.62, 95% CI 1.58–1.67), anxiety disorders (HR 2.08, 95% CI 2.03–2.14) and depressive disorders (HR 1.99, 95% CI 1.94–2.03), and with higher comorbidity (rate ratio 1.58, 95% CI 1.56–1.59) and frailty (mean ratio 1.57, 95% CI 1.56–1.58).

Life events were identified accurately from clinical notes (F1 0.81–0.95), so the low prevalences largely reflect what clinicians documented. Because linkage to population registers was not possible, we compared our estimates with national data for the events where a comparable estimate existed, and these comparisons are indicative. Financial distress was documented in 2.7% of patients in a given year, whereas 7.5% of households reported difficulties in making ends meet in national statistics (17). Violence was documented in 0.4% of patients in a given year, whereas the annual prevalence in a national survey was 16.1% in adults aged 35–54 and 6.6% in those aged 55– 74 (18). The discrepancy was large for violence, which patients may withhold out of distrust, loyalty to family members or fear of consequences such as child custody measures (19), and that clinicians may avoid asking about because of time constraints, discomfort or uncertainty about how to respond (20,21). Documentation thus reflects both how often events occur and the circumstances in which they are disclosed – and discreet events are less likely to be disclosed.

Part of the psychiatric association probably reflects how events and diagnoses are documented together, as clinicians may record an event when it supports a diagnosis. Consistent with this, excluding events documented on the day of diagnosis weakened the associations between life events and mental disorders. The physical health associations were weaker in patients with a psychiatric diagnosis (for any documented event, rate ratio for comorbidity 1.76, 95% CI 1.74–1.78 without versus 1.33, 95% CI 1.29–1.38 with a diagnosis), possibly because the diagnosis itself follows the event and carries part of the association.

Medical risk information is structured and easy to find in electronic health records, whereas psychosocial circumstances are scattered across years of clinical notes. Our findings suggest that this information is health-relevant and worth retrieving. NLP makes documented events retrievable at the patient level, which could support structured inquiry about stressful life events.

### 4.1 Limitations

Events were dated by documentation and not by occurrence, so the results describe associations with documented events, not the effects of the events themselves. Documentation is also selective, and patients with more healthcare contact have more opportunities for an event to be recorded, which may strengthen associations. Residual confounding by lifestyle behaviors remains plausible, as electronic records do not capture these variables in a structured format. Discrimination of the models for mental disorders was modest (concordance: 0.68–0.70), so documented events are better read as risk markers than as predictors. While models were internally validated, analogous data for external validation were not available.

## 5. Conclusions

Stressful life events are sparsely documented in clinical notes, but the events that are recorded are associated with mental disorders, comorbidity and frailty. Making documented life events more visible to clinicians could help them weigh psychosocial circumstances alongside medical risk information.

## Data Availability

The electronic health records analyzed in this study are not publicly available, as they contain sensitive health-related information.

## Author contributions statement

A.P. and J.J. conceived and designed the study. A.P. and J.K. developed the NLP models. A.P. and E.K. performed the statistical analyses. A.P. drafted the manuscript. J.J. supervised the study. J.J., M.V.B., and A.T. acquired the data. J.L. provided the NLP pipeline used in the study. N.A. contributed to the design of the models for psychiatric outcomes. All authors interpreted the results and approved the final manuscript.

## Ethical approval and consent to participate

In line with Finnish legislation (Act on Secondary Use of Health and Social Data 552/2019), the Human Sciences Ethics Committee of the University of Jyväskylä waived ethical review and informed consent for this study. A research permit was obtained from the Central Finland Wellbeing Services County.

