## Supplementary tables for "Stressful life events recorded in 39.7 million clinical notes: associations with mental disorders, comorbidity, and frailty"

### eTable 1. Most frequent expressions for each life event category

The 30 most frequent expressions identified for each category, in descending order of frequency, as extracted from the clinical notes, with preliminary English translations. Expressions are shown as extracted, including fragments of longer phrases, inflected forms and misspellings.

| Expression | English translation |
|---|---|
| **Grief** | |
| isä kuollut | father deceased |
| äiti kuollut | mother deceased |
| puoliso kuollut | spouse deceased |
| leskeksi | (became) a widow(er) |
| leski | widow(er) |
| veli kuollut | brother deceased |
| mies kuollut | husband deceased |
| <nimi> kuollut | [name] deceased |
| vaimo kuollut | wife deceased |
| leskeytynyt | widowed |
| äidin kuolema | mother's death |
| tehnyt itsemurhan | died by suicide |
| puoliso on kuollut | spouse has died |
| poika kuollut | son deceased |
| miehen kuolema | husband's death |
| isä kuoli | father died |
| menehtynyt | passed away |
| isä on kuollut | father has died |
| sisko kuollut | sister deceased |
| tytär kuollut | daughter deceased |
| sisar kuollut | sister deceased |
| isän kuolema | father's death |
| menehtyminen | passing away |
| äiti on kuollut | mother has died |
| leskenä | as a widow(er) |
| kuolleet | deceased |
| äiti kuoli | mother died |
| puolison kuolema | spouse's death |
| puoliso kuoli | spouse died |
| edesmennyt | the late, deceased |
| **Divorce** | |
| eronnut | divorced, separated |
| ex-puoliso | ex-spouse |
| ex-vaimo | ex-wife |
| ex-mies | ex-husband |
| eronneet | divorced, separated |
| eron jälkeen | after the divorce or separation |
| ex-puolison | ex-spouse's |

| Expression | English translation |
| --- | --- |
| eroprosessi | divorce/separation process |
| tullut ero | divorce/separation occurred |
| entinen puoliso | former spouse |
| aiemmasta liitosta | from a previous union |
| avioeron | of the divorce |
| ex-miehen | ex-husband's |
| ex-vaimon | ex-wife's |
| ex-puolisolle | to the ex-spouse |
| avioero | divorce |
| entinen mies | former husband |
| entinen vaimo | former wife |
| päättynyt eroon | ended in divorce/separation |
| ex-puolisoon | to the ex-spouse |
| aiemmasta avioliitosta | from a previous marriage |
| ex-puolisonsa | their ex-spouse |
| edellisestä liitosta | from the previous marriage |
| tuli ero | a separation occurred |
| päättyi eroon | ended in divorce or separation |
| parisuhde kariutunut | relationship has broken down |
| ex-puolisolla | at the ex-spouse's |
| parisuhde päättynyt | relationship has ended |
| ex-vaimonsa | his ex-wife |
| aiemmasta suhteesta | from a previous relationship |
| **Financial distress** | |
| työttömänä | (as) unemployed |
| toimeentulotukea | social assistance [income support] |
| toimeentulotuen | of social assistance |
| ole rahaa | (has) no money |
| työttömäksi | (became) unemployed |
| toimeentulotukihakemus | social assistance application |
| työtön | unemployed |
| ole varaa | cannot afford |
| velkaa | debt |
| toimeentulotuki | social assistance |
| toimeentulotukeen | to social assistance |
| ollut rahaa | (has not) had money |
| maksamatta | unpaid, without paying |
| työttömyyden | of unemployment |
| velkoja | debts |
| taloudellisen tilanteen vuoksi | due to the financial situation |
| ei rahaa | no money |
| irtisanottu | dismissed (from employment) |
| toimeentulotukihakemuksen | of the social assistance application |

| Expression | English translation |
|---|---|
| ulosottoon | to debt enforcement |
| taloudellisia huolia | financial worries |
| työttömyys | unemployment |
| lomautettuna | on temporary layoff |
| toimeentulotuesta | from social assistance |
| taloudelliset huolet | financial worries |
| rahaton | penniless |
| taloudellisia vaikeuksia | financial difficulties |
| rahahuolia | money worries |
| rahahuolet | money worries |
| rahaa ei ole | there is no money |
| **Violence** | |
| pahoinpidelty | assaulted |
| pahoinpitely | assault |
| väkivalta | violence |
| hyväksikäyttöä | abuse, exploitation |
| puukotettu | stabbed |
| väkivaltaa | violence |
| hyväksikäyttö | abuse, exploitation |
| pahoinpitelyn | of the assault |
| pahoinpidellyt | assaulted |
| puukotus | stabbing |
| hyväksikäytetty | abused, exploited |
| väkivallan | of violence |
| pahoinpitelystä | of assault |
| kuristettu | strangled |
| seksuaalisesti hyväkseen | sexually exploited |
| seksuaalisesti hyväksi | sexually exploited |
| hyväksikäyttänyt | (has) abused, exploited |
| väkivaltainen | violent |
| pahoinpidellyksi | (to be) assaulted |
| puukotuksen | of the stabbing |
| pahoinpitelyä | assault |
| turvakotiin | to a domestic violence shelter |
| kuristi | strangled |
| lähisuhdeväkivalta | intimate partner violence |
| kaltoinkohtelu | maltreatment |
| puukotti | stabbed |
| perheväkivalta | domestic violence |
| kaltoinkohdeltu | maltreated |
| pahoinpideltiin | was assaulted |
| pahoinpitelyyn | to assault |
| **Family problems** | |

| Expression | English translation |
|---|---|
| perhetyöntekijä | family worker |
| lastensuojelu | child welfare services (child protective services) |
| perhetyö | family work (family support services) |
| lastensuojelun | of child welfare services |
| huostaanotettu | (child) taken into care |
| tukiperhe | support family |
| otettu huostaan | (child) taken into care |
| huostaanotto | taking into care |
| huoltajuuskista | custody dispute |
| perhetyöntekijän | family worker's |
| perhetyön | of family work |
| perheterapeutti | family therapist |
| lastensuojeluilmoitus | child welfare notification |
| lastensuojeluilmoituksen | of the child welfare notification |
| perhekriisi | family crisis |
| lastensuojelusta | from child welfare services |
| lastensuojeluun | to child welfare services |
| perhetyötä | family work |
| huostaanoton | of the taking into care |
| sijaisperheessä | in a foster family |
| hankala perhetilanne | difficult family situation |
| ls-ilmoitus | child welfare notification |
| nuorisokoti | youth home |
| vaikea perhetilanne | difficult family situation |
| tukiperhettä | support family |
| poika sijoitettu | son placed (in care) |
| sijaisperhe | foster family |
| tukiperheessä | in a support family |
| lapset sijoitettu | children placed (in care) |
| perhetyöntekijät | family workers |

### eTable 2. Univariable associations between life events and psychiatric outcomes

Results from time-varying Cox proportional hazards models with age as the time scale. Each life event and the sum of life events were modelled separately, adjusted for sex only. Abbreviations: HR = hazard ratio; CI = 95% confidence interval.

| | Stress-related disorders *n = 1,617,756* *N cases = 4,481* | | Anxiety disorders *n = 1,623,606* *N cases = 3,939* | | Depressive disorders *n = 1,595,573* *N cases = 6,267* | |
|---|---|---|---|---|---|---|
| **Life event** | **HR (95% CI)** | **p** | **HR (95% CI)** | **p** | **HR (95% CI)** | **p** |
| Grief | 2.66 (2.49–2.84) | <0.001 | 3.36 (3.14–3.60) | <0.001 | 2.73 (2.58–2.89) | <0.001 |
| Divorce | 2.93 (2.68–3.22) | <0.001 | 5.18 (4.77–5.63) | <0.001 | 4.79 (4.45–5.15) | <0.001 |
| Financial distress | 1.89 (1.75–2.05) | <0.001 | 3.86 (3.60–4.15) | <0.001 | 3.41 (3.21–3.63) | <0.001 |
| Violence | 2.45 (2.11–2.84) | <0.001 | 4.81 (4.25–5.43) | <0.001 | 3.40 (3.01–3.84) | <0.001 |
| Family problems | 3.10 (2.48–3.87) | <0.001 | 4.65 (3.77–5.72) | <0.001 | 3.41 (2.74–4.25) | <0.001 |
| *Sum of life events* | *1.63 (1.58–1.67)* | *<0.001* | *2.08 (2.03–2.14)* | *<0.001* | *1.99 (1.94–2.03)* | *<0.001* |

### eTable 3. Concordance of the models for psychiatric outcomes

Concordance of the models in Table 4. Abbreviations: SE = standard error.

| | Stress-related disorders | Anxiety disorders | Depressive disorders |
|---|---|---|---|
| **Model** | ***Concordance*** | ***Concordance*** | ***Concordance*** |
| Individual life events | 0.683 (0.004) | 0.704 (0.005) | 0.677 (0.004) |
| *Sum of life events* | *0.681 (0.004)* | *0.704 (0.005)* | *0.677 (0.004)* |

### eTable 4. Concordance of the models for psychiatric outcomes in training and test data

Concordance of the models in Table 4, estimated in a random 70/30 split of participants into training and test sets, with the models fitted in the training set.

| | Stress-related disorders | | Anxiety disorders | | Depressive disorders | |
|---|---|---|---|---|---|---|
| | ***Concordance*** | | ***Concordance*** | | ***Concordance*** | |
| **Model** | ***Train*** | ***Test*** | ***Train*** | ***Test*** | ***Train*** | ***Test*** |
| Individual life events | 0.691 | 0.670 | 0.708 | 0.694 | 0.677 | 0.680 |
| *Sum of life events* | *0.687* | *0.666* | *0.706* | *0.700* | *0.676* | *0.678* |

### eTable 5. Sensitivity analysis additionally excluding individuals with life events documented on the date of diagnosis

Results from Cox proportional hazards models with age as the time scale, adjusted for sex and baseline CCI. Individuals whose life event was documented on or after the date of diagnosis in the year of diagnosis were excluded (the main analysis in Table 4 excluded only those documented after the date of diagnosis). The five life event categories are mutually adjusted; the sum of life events was estimated in a separate model. HR = hazard ratio; CI = 95% confidence interval.

| | **Stress-related disorders** *No. of cases = 3,904* | | **Anxiety disorders** *No. of cases = 3,633* | | **Depressive disorders** *No. of cases = 5,713* | |
|---|---|---|---|---|---|---|
| **Life event** | **HR (95% CI)** | **p** | **HR (95% CI)** | **p** | **HR (95% CI)** | **p** |
| Grief | 1.53 (1.41–1.66) | <0.001 | 2.05 (1.89–2.22) | <0.001 | 1.75 (1.64–1.87) | <0.001 |
| Divorce | 1.89 (1.66–2.14) | <0.001 | 2.38 (2.13–2.67) | <0.001 | 2.35 (2.13–2.60) | <0.001 |
| Financial distress | 1.22 (1.09–1.35) | <0.001 | 2.13 (1.94–2.34) | <0.001 | 2.06 (1.90–2.22) | <0.001 |
| Violence | 1.13 (0.93–1.37) | 0.233 | 1.64 (1.40–1.92) | <0.001 | 1.45 (1.25–1.68) | <0.001 |
| Family problems | 1.48 (1.13–1.95) | 0.005 | 1.18 (0.92–1.51) | 0.195 | 1.08 (0.84–1.39) | 0.562 |
| *Sum of life events* | *1.45 (1.40–1.50)* | *<0.001* | *2.01 (1.96–2.07)* | *<0.001* | *1.87 (1.82–1.92)* | *<0.001* |

### eTable 6. Univariable associations between life events and trajectories of comorbidity and frailty

Results from generalized linear mixed models with a random intercept and a random slope for age, fitted as Poisson for comorbidity and Tweedie for frailty. Comorbidity was measured with the Charlson comorbidity index (CCI) and frailty with the Hospital frailty risk score (HFRS). Each life event and the sum of life events were modelled separately, adjusted for sex and centered age only. Each model included 1,658,163 observations. CI = 95% confidence interval.

| | **Comorbidity** | | **Frailty** | |
|---|---|---|---|---|
| **Life event** | **Rate ratio (95% CI)** | **p** | **Mean ratio (95% CI)** | **p** |
| Grief | 1.57 (1.55–1.59) | <0.001 | 1.60 (1.58–1.61) | <0.001 |
| Divorce | 1.79 (1.75–1.84) | <0.001 | 1.85 (1.82–1.88) | <0.001 |
| Financial distress | 1.89 (1.85–1.92) | <0.001 | 1.86 (1.84–1.89) | <0.001 |
| Violence | 1.59 (1.52–1.66) | <0.001 | 1.62 (1.58–1.66) | <0.001 |
| Family problems | 1.72 (1.58–1.88) | <0.001 | 1.85 (1.76–1.95) | <0.001 |
| *Sum of life events* | *1.52 (1.51–1.54)* | *<0.001* | *1.56 (1.55–1.57)* | *<0.001* |

### eTable 7. Performance of the mixed models for comorbidity and frailty

Marginal $R^2$ = variance explained by fixed effects; conditional $R^2$ = variance explained by fixed and random effects. Akaike Information Criterion (AIC) and Bayesian Information Criterion (BIC) are comparable only between models of the same outcome (the comorbidity and frailty models use different distributions). SD = standard deviation.

| | Comorbidity | | Frailty | |
|---|---|---|---|---|
| **Metric** | **Individual events** | **Sum of life events** | **Individual events** | **Sum of life events** |
| Marginal $R^2$ | 0.378 | 0.378 | 0.394 | 0.394 |
| Conditional $R^2$ | 0.998 | 0.998 | 0.996 | 0.996 |
| AIC | 2,018,578 | 2,018,781 | 4,599,030 | 4,599,322 |
| BIC | 2,018,726 | 2,018,880 | 4,599,203 | 4,599,445 |
| Random intercept, SD | 2.760 | 2.765 | 2.898 | 2.901 |
| Random slope, SD (age) | 0.108 | 0.108 | 0.104 | 0.104 |
| Intercept–slope correlation | −0.34 | −0.34 | −0.29 | −0.28 |
